# Risk factors and survival in patients with COVID-19 in northeastern Brazil

**DOI:** 10.1101/2022.01.27.22269602

**Authors:** Ana Tereza do N S F Fernandes, Eujessika K. Rodrigues, Eder R. Araújo, Magno F. Formiga, Priscilla K. Sá Horan, Ana Beatriz N. S. Ferreira, Humberto A. Barbosa, Paulo S. Barbosa

## Abstract

**BACKGROUND:** Countries have focused research on developing strategies to fight COVID-19, prevent hospitalizations, and maintain economic activities.

**OBJECTIVE:** This study aimed to establish a survival analysis and identify risk factors for patients with COVID-19 in a upper middle-income city in Brazil.

**METHODS:** We performed a retrospective cohort study with 280 hospitalized patients with COVID-19. The eCOVID platform provided data used to monitor COVID-19 cases and help communication between professionals.

**RESULTS:** Survival analysis showed that age ≥ 65 years was associated with decreased survival (54.8%). Females had lower survival rate than males (p=0.01). Regarding risk factors, urea concentration (p<0.001), hospital LOS (p=0.002), oxygen concentration (p=0.005), and age (p=0.02) were associated with death.

**CONCLUSION:** Age, hospital LOS, high blood urea concentration, and low oxygen concentration were associated with death by COVID-19 in the studied population. These findings corroborate with studies conducted in research centers worldwide.

**Key Findings:** Some parameters assessed during hospital admission may identify patients with COVID-19 with high risk of progressing to severe conditions and early identification of risk factors and continuous monitoring of laboratory tests may prevent progression to severe disease.

**Key Implications:** Knowledge regarding main signs and symptoms of COVID-19 and associated risk factors helps the population manage and monitor the disease at home and identify early signs of severity. Also, more information about COVID-19, such as prevalence, specific characteristics in certain regions, and treatments, must become public to encourage the population to participate in the control and contingency of the pandemic.

## Introduction

A transmissible disease (COVID-19) caused by the SARS-CoV-2 virus increased the number of severe acute respiratory syndrome (SARS) cases worldwide since December 2019. Although Efforts were made to determine prevalence and factors associated with severity of COVID-19^1,2^. To date, more than 224 million people have been infected, and more than four million died. The United States of America and Brazil correspond to 71% of cumulative cases of COVID-19 in the Americas (more than 46 million cases)^3^.

Many countries focused their research on defining and identifying strategies to fight the disease, prevent hospitalizations, and maintain economic activities^4–6^. One critical point of the disease is its form of presentation, varying from asymptomatic and very mild to critical symptoms. Symptoms may also persist even after the acute phase, and individuals who initially did not report or had mild symptoms may evolve to sudden health decline or death^4,7,8^.

Identifying factors that predispose to high risk of hospitalization and death contributes to preventive measures, especially in developing countries facing difficulties in establishing early diagnosis^8^. Therefore, this study describes clinical and demographic characteristics, comorbidities, outcomes, survival and factors associated with mortality of hospitalized patients with COVID-19 in an upper-middle-income city of Brazil.

## Methods

### Study design and participants

A retrospective cohort study was conducted with patients hospitalized due to COVID-19. The study followed the Strengthening the Reporting of Observational Studies in Epidemiology (STROBE)^1^.

Data were collected from the virtual platform eCOVID^2^, developed by the Center for Strategic Health Technologies of the State University of Paraíba (NUTES/UEPB) in Brazil. Qualified professionals obtained data during admission of patients in two hospitals of the city. The study was approved by the local research ethics committee (approval number 4.241.971) and followed all ethical aspects involving research in humans and the Declaration of Helsinki.

We included hospitalized patients of both sexes, aged 18 or over, diagnosed with COVID-19 using RT-PCR test (nasopharynx swab)^3^, with epidemiological history of COVID-19 and/or relevant clinical symptoms and serological parameters^9,10^. Patients whose COVID-19 diagnosis was discarded or outcomes were not recorded were excluded.

### eCOVID platform

The eCOVID platform was developed to store patient data (e.g., symptoms, comorbidities, vital signs, and laboratory and imaging exams) and facilitate communication among professionals treating patients with COVID-19. The platform allows remote assistance/communication among professionals working in the public Brazilian Unified Health System and the private sector. It also provides a database for epidemiological studies in the city where the study was conducted.

### Variables and data collection

The following variables were included for data analysis: (1) profile of patients (age and sex); (2) diagnostic method (RT-PCR, immunoglobulin serology, or clinical-epidemiological); (3) symptoms presented on admission; (4) comorbidities; (5) vital signs; (6) laboratory and imaging exams; (7) risk stratification on admission; (8) hospital length of stay (LOS); (9) length of invasive mechanical ventilation (IMV); and (10) clinical outcomes (discharge or death). We obtained laboratory parameters, such as blood cell count and renal function (i.e., concentration of urea and creatinine), from medical records.

Data from the first day (admission) to the last day of hospitalization (hospital discharge/death) were included for analysis. Vital signs, symptoms, and laboratory tests were considered if obtained on admission, while data regarding IMV, hospital LOS, and imaging tests were obtained when the outcome (discharge or death) was recorded.

### Data Analysis

The multiple imputation approach (intention to treat analysis)^11^ estimated plausible values and replaced missing data from laboratory tests. We computed five different imputations and generated five individual databases and five results. Then, results were grouped, and inferences were performed. The multiple imputation approach generated more accurate values than those generated by single imputation methods and also helped to understand patterns of missing data from our data set.

Categorical data were presented as relative frequency and numerical data as mean and standard error (SE). Patients were divided into survivors (patients discharged) and non-survivors (patients who died). Kolmogorov-Smirnov test assessed data normality, and the unpaired t-test compared data between groups. Chi-square test (Chi^2^) compared comorbidities, signs, and symptoms between groups. Cox (proportional hazards) regression investigated the effects of age (< 65 and ≥ 65 years) and sex on death, whereas survival analysis was performed using Kaplan-Meier method. We also developed logistic regression models using the progressive selection technique (i.e., forward selection), in which only significant variables (p ≤ 0.05) and predictive value remained in the model to identify those interfering with death. studied outcomes. The software Statistical Package for the Social Sciences version 22 (IBM Corp., USA) performed inferential analysis, and significance was set at p ≤ 0.05.

## Results

### Clinical Characteristics

Table 1 shows general and clinical characteristics, symptoms, and comorbidities presented by the studied population. Among 280 hospitalized patients with COVID-19 included in our study, 192 were discharged, and 88 died. Approximately 54% of patients were males with mean age of 62.42 years (SE: 1.45). The most common comorbidities were systemic arterial hypertension (SAH) (53%), diabetes mellitus (67%), and obesity (24%). Moreover, 10% of patients had other types of heart disease. Dyspnea was the most reported symptom (82%), followed by cough (59%), fever (57%), muscular pain (22%), headache (15%), and anosmia (11%). Furthermore, 27% of patients were critically ill, and 57% were moderately ill on admission.

**Table 1.**
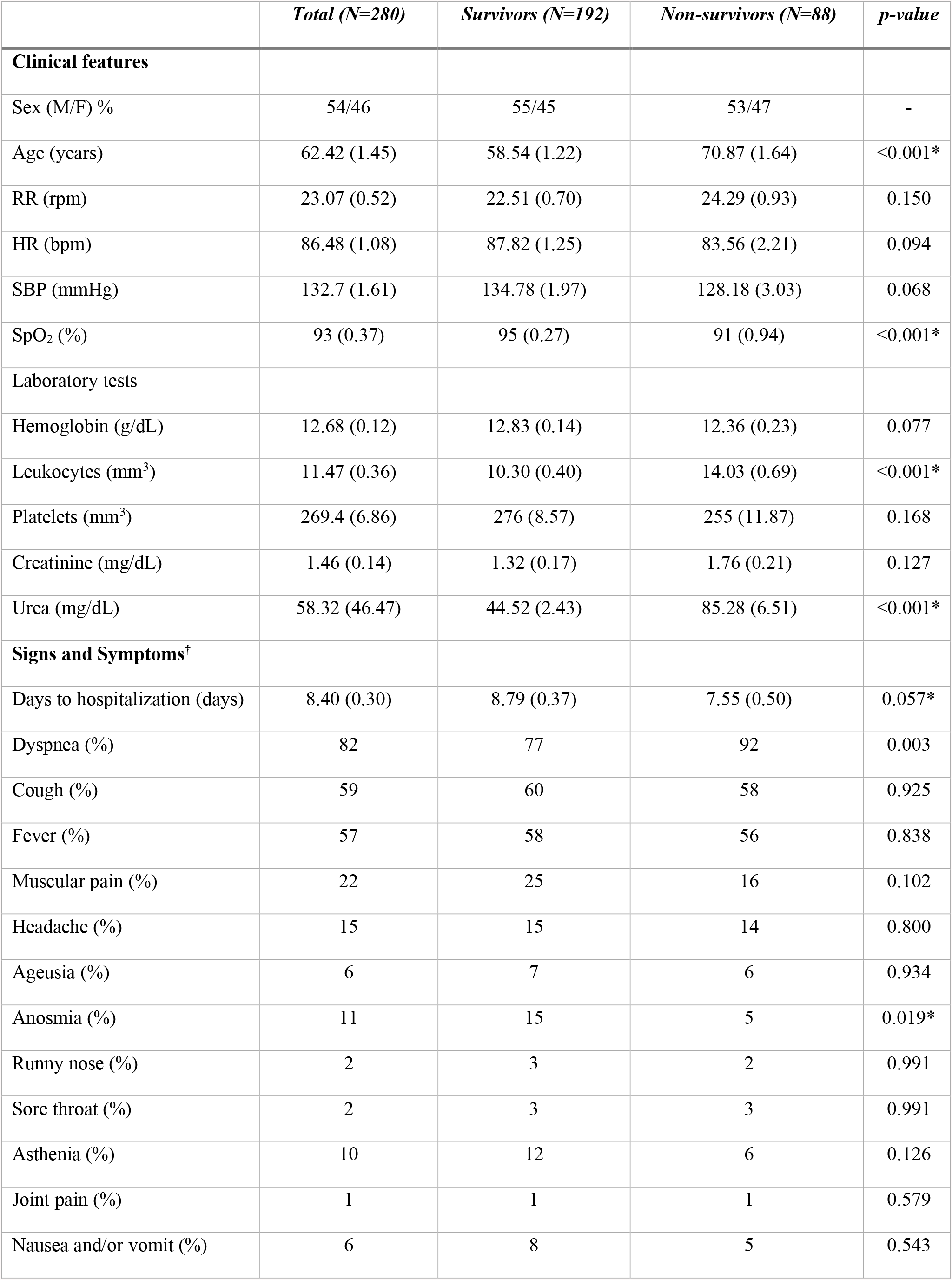
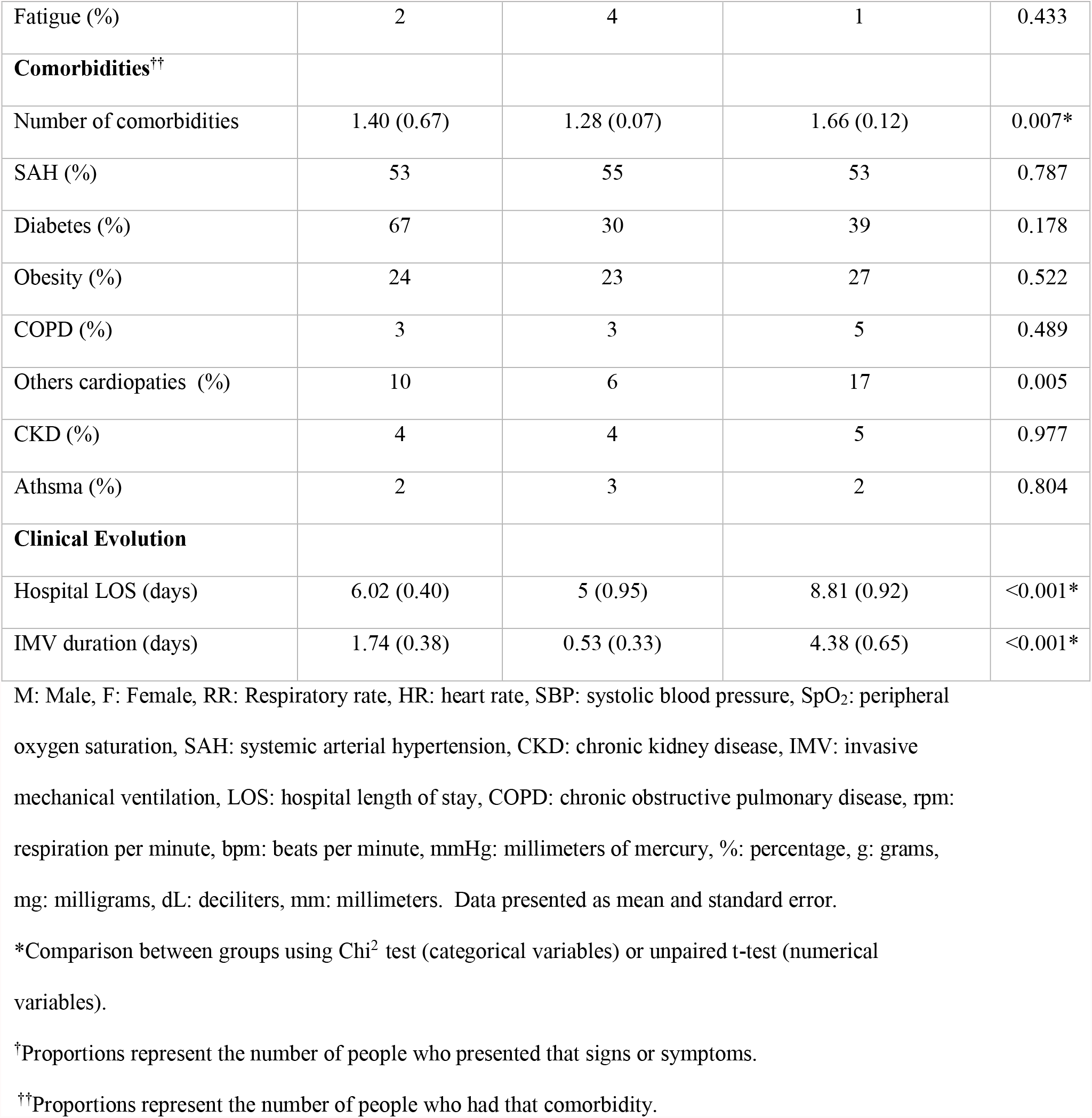
Clinical characteristics, laboratory tests, signs/symptoms, and comorbidities.

Age (p = 0.001), dyspnea (p = 0.003), and presence of other heart diseases (p = 0.005) were different between groups. SAH was the most reported comorbidity in the non-survivor group (53%). This group also presented a high prevalence of cough (58%) and fever (56%).

We observed significant differences in SpO_2_ (p < 0.001), leukocyte count (p < 0.001), blood urea concentration (p < 0.001), anosmia (p = 0.019), number of associated comorbidities (p = 0.007), hospital LOS (p < 0.001), length of IMV (p < 0.001), and time between symptom onset and hospitalization (p = 0.057).

### Survival Analysis

The overall model, including age and sex as predictors, significantly improved the fit compared with null model [χ2 (2) = 8.395, p = 0.01]. With increasing age, hazard associated with death tended to be greater, even though this predictor was not significant (β = 0.424, p = 0.07). On the other hand, sex was an independent and significant predictor of mortality hazard (β = 0.461, p = 0.03), with time until death higher for males than females.

Survival analysis showed that 81.4% of patients aged <65 years survived (Figure 1A). Survival rate in patients aged ≥ 65 years reduced to 54.8% and reached 50% in eleven days of hospitalization. In contrast, patients aged < 65 years reached a survival rate of 50% only on the 14^th^ day of hospitalization (p = 0.04). Regarding sex, survival analysis showed that 67.7% of females and 69.3% of males survived. Females and males reached 50% of survival rate on the 12^th^ and 17^th^ day of hospitalization (p = 0.01), respectively (Figure 1B).

**Figure 1.**
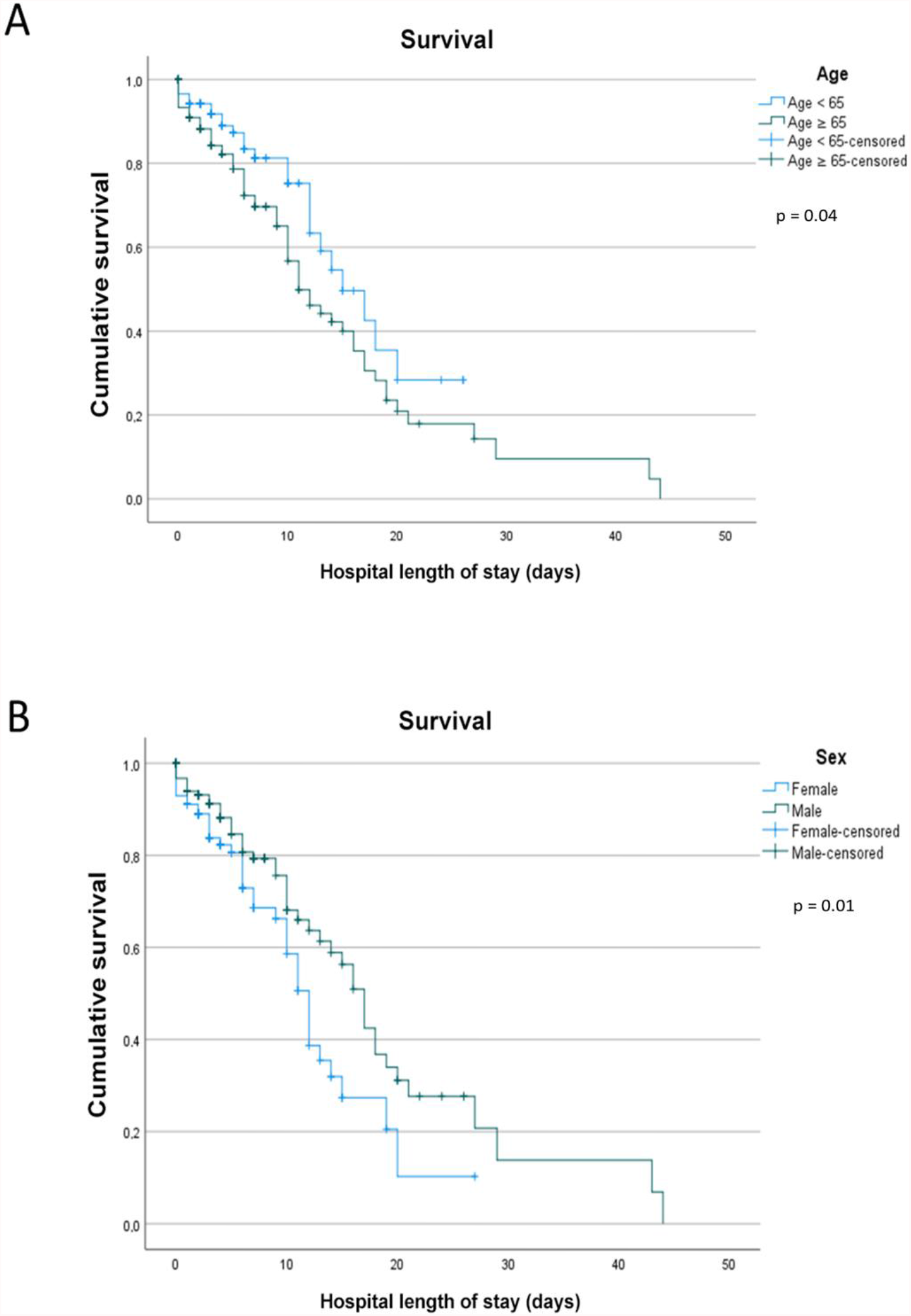
Survival analysis by (a) age and hospital length of stay and (b) sex and hospital length of stay

### Risk factors associated with mortality

The logistic regression model included all numerical variables assessed (Table 2). Urea concentration (p < 0.001), hospital LOS (p = 0.002), SpO_2_ (p = 0.005), and age (p = 0.02) were associated with death (Hosmer and Lemeshow test, p = 0.50). These variables were also different between groups.

**Table 2.** Logistic regression of variables associated with death.

|  | **RISK OF DEATH** |  |  |  |
| --- | --- | --- | --- | --- |
| | $\beta$ | p-value | OR | 95% CI |
| Age | 0.02 | 0.02 | 1.02 | 1.00-1.05 |
| SpO<sub>2</sub> | -0.11 | 0.005 | 0.89 | 0.82-0.96 |
| Hospital LOS | 0.10 | 0.002 | 1.10 | 1.03-1.18 |
| Urea | 0.03 | <0.001 | 1.03 | 1.02-1.04 |
| Constant | 5.18 | 0.15 | 177.94 |  |

OR: Odds ratio, 95% CI: confidence interval, SpO<sub>2</sub>: peripheral oxygen saturation, LOS: length of stay.

## Discussion

This study analyzed data from 280 hospitalized patients with COVID-19. Most common comorbidities were SAH, diabetes mellitus, and obesity, while most reported symptoms were dyspnea, cough, and fever. Age ≥ 65 years, low SpO_2_, high urea concentration, and prolonged hospital LOS were associated with increased risk of death in these patients. Moreover, females were associated with low survival rate when hospital LOS lasted more than eleven days.

Hospitalization due to COVID-19 is linked to worsening symptoms and increased risk of developing SARS, which increased in Brazil between 2019 and 2020^12^. Thus, identifying clinical characteristics of patients who reach severe clinical conditions is a global concern.

Casas-Rojo et al.^13^ analyzed data from 15,111 hospitalized patients of 150 hospitals in Spain and found a high prevalence of males in the sample (more than 57% aged > 60 years). These authors also observed that SAH (50.9%), obesity (21.2%), and diabetes mellitus (19.4%) were the most prevalent comorbidities, while most reported signs and symptoms on admission were fever (63.4%), dry cough (58%), and dyspnea (57.6%). Results of our study were similar to those observed in the Spanish population^13^, suggesting a pattern of clinical characteristics of hospitalized patients with more severe COVID-19. Similarly, Cabrini et al.^14^ evaluated data from 1,591 patients admitted to intensive care units in Italy and found that most hospitalized patients were males (82%) with SAH (49%). Furthermore, 89% of patients aged > 64 years needed IMV, increasing mortality rate and hospital LOS^12^. In our study, 28% of patients received IMV, and those who died needed this type of support for a longer period (4.38 days). We highlight that prolonged hospital LOS was associated with high risk of death.

Although studies showed a high prevalence of SARS-CoV-2 or other types of coronavirus in males^15,16^, survival rate dropped faster in females than males and presented a reduced percentage of survival. The survival analysis conducted by Salinas-Escudero et al.^17^ with 133 Mexican patients showed high mortality in older females, especially aged > 75 years. Our data also showed higher mortality in females than males (32.3% vs. 30.7%) but without significant difference between groups (p= 0.77), probably due to the high rate of deaths not related to sex particularities.

Studies showing age as risk factor for worsening COVID-19 are frequent in the literature^13,14,18^. Li et al.^19^ showed that age ≥ 65 years was associated with risk of severe disease and SAH. In the present study, survival rate in females aged ≥ 65 years declined to 50% when hospitalization was longer than eleven days. Also, mean age of patients who died was 70.87 years, 12 years more than those discharged. Although age as risk factor for developing COVID-19 is still controversial, its role in mortality is well established^20^. In this sense, are the aspects inherent to aging responsible for developing the disease, or is the presence of individual factors (e.g., immune response), comorbidities, and particular aspects of the elderly that worsen the response to viral infection? As already established, chronic diseases are common in the elderly, causing health^15^ problems, especially during COVID-19 infection.

Other factors, such as SpO_2_, hospital LOS, and blood urea concentration were associated with mortality in our sample. Blood urea concentration was also associated with COVID-19 severity in the study by Ok et al.^21^. These authors observed that patients who progressed to severe disease had higher urea concentration than those with moderate disease. Moreover, urea/creatinine ratio, white cell count, C-reactive protein, monocyte/lymphocyte ratio, and neutrophil/lymphocyte ratio were considered predictive factors for disease severity. These findings reinforce that early assessment may identify patients with risk of disease worsening.

Prolonged hospitalization, mainly in older individuals, was associated with mortality. Thai et al. ^18^ showed that patients who died had mean hospital LOS of eight days, whereas those who survived had hospital LOS of six days. Prolonged hospitalization causes damage in muscle strength, quality of life and functionality domains in older patients.^22^ The negative impact on physical, cognitive, and social well-being in COVID-19 patients who recover from ICU admissions and prolonged LOS due to acute respiratory illnesses is already recognized.^23^

Limited access of the population to COVID-19 diagnostic test and its acquisition and availability by the health system may be a potential limitation of this study. Specifically during hospital care, some patients were already outside the window of time to detect SARS-CoV-2 using RT-PCR. Therefore, diagnosis was performed by identifying immunoglobulins M and G and clinical and/or radiological evolution of the patient. Furthermore, the unfamiliarity of the population with signs and symptoms of COVID-19 may have delayed healthcare seeking. As a result, patients arrived at the hospital with advanced disease.

## Conclusions

Age, hospital LOS, high blood urea concentration, and low SpO_2_ are associated with mortality by COVID-19 in the evaluated population. Considering that several variables associated with increased mortality are assessed at hospital admission, this study may guide the development of public health interventions to prevent clinical evolution of COVID-19 to severe conditions. Furthermore, health professionals can assist and contribute to prevention by early identifying patients with severe COVID-19 in primary care.

## Data Availability

All data produced are available online at doi 10.6084/m9.figshare.18858473

https://figshare.com/s/439d0d45b7b3e442728f

## Fundings and Acknowledgments

This study was financed by the Coordenação de Aperfeiçoamento de Pessoal de Nível Superior – Brasil (CAPES) (Finance Code CAPES EPIDEMIAS 09/2020) and Fundação de Apoio à Pesquisa do Estado da Paraíba – Brasil (FAPESQ) (Finance Code COVID-19 003/2020). The authors thank Probatus Academic Services for providing scientific language revision and editing.

## Footnotes

^1^ www.equator-network.org/reporting-guidelines/strobe/

^2^ ecovid.nutes.uepb.edu.br

## References

1. Cao X. COVID-19: immunopathology and its implications for therapy. Nature Reviews Immunology. 2019;2019:2019–2020. doi:10.1038/s41577-020-0308-3

2. Seyed Hosseini E, Riahi Kashani N, Nikzad H, Azadbakht J, Hassani Bafrani H, Haddad Kashani H. The novel coronavirus Disease-2019 (COVID-19): Mechanism of action, detection and recent therapeutic strategies. Virology. 2020;551(September):1–9. doi:10.1016/j.virol.2020.08.011

3. WHO | World Health Organization. Accessed May 13, 2021. https://www.who.int/

4. Machhi J, Herskovitz J, Senan AM, et al. The Natural History, Pathobiology, and Clinical Manifestations of SARS-CoV-2 Infections. Published online 2020.

5. Wang D, Hu B, Hu C, et al. Clinical Characteristics of 138 Hospitalized Patients with 2019 Novel Coronavirus-Infected Pneumonia in Wuhan, China. JAMA - Journal of the American Medical Association. 2020;323(11):1061–1069. doi:10.1001/jama.2020.1585

6. Stawicki SP, Jeanmonod R, Miller AC, et al. The 2019 – 2020 Novel Coronavirus (Severe Acute Respiratory Syndrome Coronavirus 2) Pandemic : A Joint American College of Academic International Medicine-World Academic Council of Emergency Medicine Multidisciplinary COVID-19 Working Group Consensus Pa. 2021;12(2):47–93. doi:10.4103/jgid.jgid

7. Borah P, Deb PK, Chandrasekaran B, Goyal M. Neurological Consequences of SARS-CoV-2 Infection and Concurrence of Treatment-Induced Neuropsychiatric Adverse Events in COVID-19 Patients : Navigating the Uncharted. 2021;8(February):1–17. doi:10.3389/fmolb.2021.627723

8. Zhou F, Yu T, Du R, et al. Since January 2020 Elsevier has created a COVID-19 resource centre with free information in English and Mandarin on the novel coronavirus COVID-research that is available on the COVID-19 resource centre - including this for unrestricted research re-use a. 2020;(January).

9. Dramé, M; Teguo, MT; Proye, E; Hequet, F; Hentzien, M; Kanagaratnam, L; Godaert L. Should RT-PCR be considered a gold standard in the diagnosis of COVID-19_ _ Enhanced Reader.pdf.

10. Wang Z, Ji JS, Liu Y, et al. Survival analysis of hospital length of stay of novel coronavirus (COVID-19) pneumonia patients in Sichuan, China. medRxiv. Published online 2020.

11. Newell Newell D J DJ. Intention-to-Treat Analysis: Implications for Quantitative and Qualitative Research. Vol 21.; 1992.

12. Luis J, Rocha L, Waib LF, Carrilho CM, Margareth S, Lobo A. Orientações sobre Diagnóstico, Tratamento e Isolamento de Pacientes com COVID-19. 2020;9(2).

13. J.M. Casas-Rojo, J.M. Antón-Santosa, *JMNC, C. Lumbreras-Bermejo, J.M. Ramos-Rincónd, E. Roy-Vallejo e AAM, F. Arnalich-Fernándezg, J.M. García-Brũnénh, J.A. Vargas-Nú∼nezi SJFC, et al. Revista Clínica Española Características clínicas de los pacientes hospitalizados con COVID-19 en Espa ∼. 2020;220(8). doi:10.1016/j.rce.2020.07.003

14. Cabrini L, Castelli A, Cereda D, Coluccello A, Foti G. Baseline Characteristics and Outcomes of 1591 Patients Infected With SARS-CoV-2 Admitted to ICUs of the Lombardy Region, Italy. 2021;323(16):1574–1581. doi:10.1001/jama.2020.5394

15. Badawi A, Ryoo SG. Prevalence of comorbidities in the Middle East respiratory syndrome coronavirus (MERS-CoV): a systematic review and meta-analysis. International Journal of Infectious Diseases. 2016;49:129–133. doi:10.1016/j.ijid.2016.06.015

16. Channappanavar R, Fett C, Mack M, et al. Sex-based differences in susceptibility to SARS-CoV infection. 2018;198(10):319–335. doi:10.4049/jimmunol.1601896.Sex-based

17. Salinas-Escudero G, Carrillo-Vega MF, Granados-García V, Martínez-Valverde S, Toledano-Toledano F, Garduño-Espinosa J. A survival analysis of COVID-19 in the Mexican population. BMC Public Health. 2020;20(1):1616. doi:10.1186/s12889-020-09721-2

18. Thai PQ, Thi D, Toan T, Son DT, Van HTH. Factors associated with the duration of hospitalisation among COVID-19 patients in Vietnam : A survival analysis. Published online 2020.

19. Li X, Xu S, Yu M, Wang K, Tao Y, Zhou Y. Risk factors for severity and mortality in adult COVID-19 inpatients in Wuhan. Journal of Allergy and Clinical Immunology. 2020;146(1):110–118. doi:10.1016/j.jaci.2020.04.006

20. Triggle CR, Bansal D, Ding H, et al. A Comprehensive Review of Viral Characteristics, Transmission, Pathophysiology, Immune Response, and Management of SARS-CoV-2 and COVID-19 as a Basis for Controlling the Pandemic. Frontiers in Immunology. 2021;12(February):1–23. doi:10.3389/fimmu.2021.631139

21. Ok F, Erdogan O, Durmus E, Carkci S, Canik A. Predictive values of blood urea nitrogen/creatinine ratio and other routine blood parameters on disease severity and survival of COVID-19 patients. Journal of Medical Virology. 2021;93(2):786–793. doi:10.1002/jmv.26300

22. Meira D, Lavoura P, Ferreira D, et al. Impact of hospitalization in the functionality and quality of life of adults and elderlies. European Respiratory Journal. 2015;46(suppl 59):PA3547. doi:10.1183/13993003.congress-2015.PA3547

23. Simpson R, Robinson L. Rehabilitation after critical illness in people with COVID-19 infection. American Journal of Physical Medicine and Rehabilitation. 2020;99(6):470–474. doi:10.1097/PHM.0000000000001443

